# Willingness to pay tuition and risk-taking proclivities among students: A fundamental conundrum for universities

**DOI:** 10.1101/2020.08.26.20182352

**Authors:** Zafar Zafari, Lee Goldman, Katia Kovrizhkin, Peter Muennig

## Abstract

**Importance:** As universities around the world decide whether to remain open or to close their campuses because of the COVID-19 pandemic, they often are doing so without objective information on the preferences and risk tolerance of their students.

**Objectives:** To quantify students’: 1) risk tolerance for in-person instruction; 2) willingness to pay for in-person instruction versus online-only instruction; and 3) risk-tolerance for social activities held off campus.

**Design, Setting, and Participants:** We developed an automated survey tool that administered a “standard gamble” exercise grounded in game theory to 46 Columbia University public health graduate students who were knowledgeable about COVID-19 and who had experience with both online and offline coursework. Students were asked to trade between the risk of becoming infected with COVID-19 and: 1) attending classes in-person versus online and 2) attending parties in the greater New York City area. We also assessed their willingness to pay for online only tuition, and plans to travel off campus.

**Main Outcome Measures:** The decision point in iterative trade-offs between risk of infection with COVID-19 and a desired goal (taking classes in-person or attending social events).

**Results:** On average, students were willing to accept a 23% (standard error [SE]: 4%) risk of infection on campus over the semester in exchange for the opportunity to attend classes in-person. Students were willing-to-pay only 48% (SE: 3%) of typical in-person tuition were courses held exclusively online, and no students were willing to pay full price for online-only instruction. Students planned to leave campus an average of 3.6 times per week (SE: 0.54), and 15% of the students would be willing to attend a party in the community surrounding the university even if the prevalence of circulating COVID-19 were high.

**Conclusions and Relevance:** Students with a strong knowledge of COVID-19 transmission and risks are an enigma: they are willing to pay only around 50% for online classes but likely to engage in activities that present significant barriers to holding in-person classes This enigma underscores the conundrum facing universities.

## Introduction

A large number of universities are open to live instruction in the Fall of 2020.^1^ These universities will open with extensive preventive measures in place but a substantial proportion of students might not adhere to them. As a result, universities face a substantial challenge. On one hand, if they re-open, students could prove to be a major pathway for the spread of COVID-19, thereby worsening the pandemic and requiring the university to suspend in-person activities. On the other hand, if universities open to online-only instruction, they will find it difficult to sustain their educational mission, maintain enrollment, and balance their budgets.^2^

Our objective was to inform university decision-makers regarding the trade-offs that students are willing to make in exchange for having classes held in-person as well as their perceived acceptability of holding classes online only. To assess students’ preferences and likely behaviors, we used a “standard gamble” exercise that assesses how individuals try to make rational decisions under uncertainty.^3^ Based on game theory, the process asks participants choose between two alternative options, each with its own probability, until they find each option to be equivalent. This tool allows a more accurate estimate of students’ preferences for risk than a simple survey alone.^4^

If students are not willing to tolerate any substantial risk of infection with SARS-CoV-2 and simultaneously are willing to pay regular tuition for online classes, then online-only instruction is a logical choice. Online-only instruction also makes sense from a public health standpoint if students are eager to socialize even in the context of a dangerous pandemic. Conversely, in-person classes would be recommended if students are unwilling to pay adequate tuition for on-line only instruction and are willing to observe public health guidelines.

For this exercise, we asked students to make a gamble between the risk COVID-19 infection in exchange for taking classes in-person. If the student is willing to take the gamble of being infected, then the stakes are raised until the student is ambivalent about the decision between online and in-person classes. Similarly, we asked students to make a trade-off between risking infection with COVID-19 and attending a party. We also assessed students’ willingness to pay for online-only instruction and the likelihood that they would take public transportation or eat out at restaurants.

## Methods

### Sample

The exercise that we deployed required underlying knowledge of the epidemiology of COVID-19 as well as experience with both in-person and online learning.^4 5^ We therefore chose to survey students who had recently completed a Master’s of Public Health program at Columbia University’s Mailman School of Public Health and who had attended both in-person classes during the Fall 2019 and early Spring 2020 semesters as well as online classes after the lockdown in the middle of the Spring 2020 semester. Administrators in the Department of Health Policy and Management at the Mailman School of Public Health identified 30 students who had graduated in Spring 2020 and had been engaged in student groups, helped with interviewing incoming students, or worked as teaching assistants.

All 30 students agreed to participate. Some of these students subsequently recruited 16 additional students to participate, resulting in a total sample size of 46 students. To maintain anonymity and expedite IRB approval, no demographic details were obtained from the students. The first 30 responses to the survey are very likely to be the first 30 students contacted as the request from students for recruiting other students came after the first 30 responses were received. We conducted t-tests to compare the means of the first 30 responses with those from the additional 16 students to ensure that the mean responses and variance did not differ from the initial 30 students.

### Survey

We used Survey Sparrow, Inc. (Palo Alto, CA) to administer the survey. Survey Sparrow deploys an interactive chat bot that implements skip logic and can accept conditional terms. Thus, it is possible to ask a subsequent question that is conditional on the response to the first question.

To assess students’ preferences and likely behaviors, we used a “standard gamble” exercise to understand how individuals try to make rational decisions under uncertainty.^3^ Based on game theory, the process asks participants choose between two alternative options, each with its own probability, until they find each option to be equivalent. This tool allows a more accurate estimate of students’ preferences for risk than a simple survey alone.^4^

Students were first presented with data on their age-specific risk of illness, hospitalization, and death as well as possible symptoms associated with COVID-19: “As a 20-25-year-old…you will typically have a flu-like illness in which you are in bed with a fever and cough. You will be quarantined for 2-3 weeks depending on the duration of your illness. If you become infected, you have a 1.04% chance of being hospitalized and a chance of requiring ventilation or dying (0.03%).” These risks were computed from a recent study that provided age-specific infection mortality rates.^6^

Students were then presented with a “standard gamble” exercise, in which they were asked to trade between the risk of illness presented from in-person instruction and the safety of online instruction (**Figure 1**). Questions regarding risk offered ‘Yes’ and ‘No’ answer options when asking students whether they were willing to accept a certain risk to take classes in-person rather than online. The gamble began with a trade-off between a 10% chance of becoming infected while attending in-person classes versus a 0% chance of becoming infected taking online classes. The bot then increased or decreased their chances of infection depending on their response until an equilibrium was reached. If the student accepted this risk, they were presented with a 5% increase in risk and asked again if they would accept this risk of infection to attend classes in-person. This exercise continued (to 100%), with risk being increased 5% with each question, until the student felt they no longer would accept the risk of infection in exchange for in-person class. If the student rejected the 10% risk, the student was presented with 5%, 1%, 0.5%, 0.1%, 0.05%, and 0.001% risk until they were willing to accept the risk. If the student rejected all presented risk, it was recorded as an acceptance of no risk of infection in exchange for attending in-person class.

**Figure 1.**
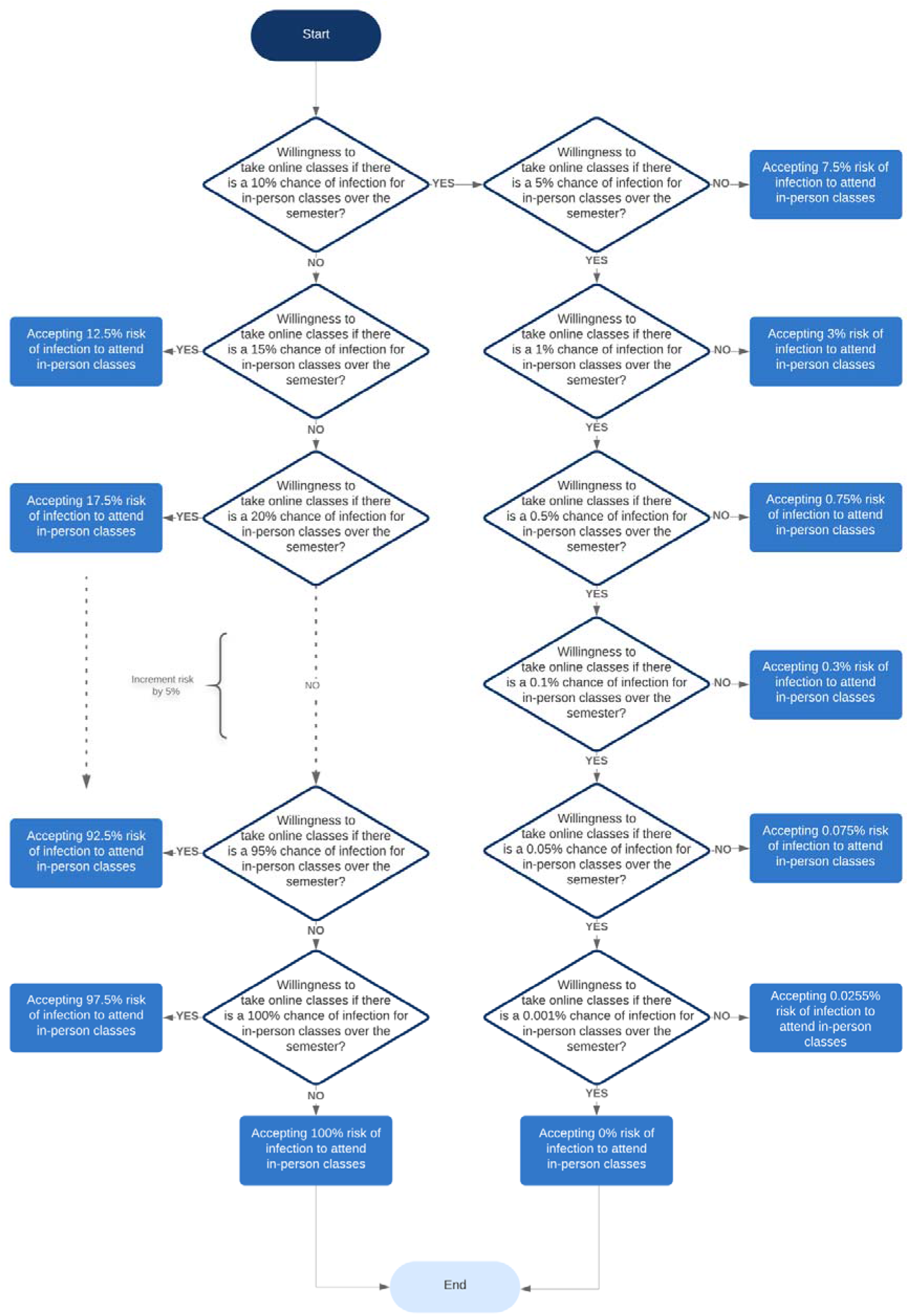
Flow diagram depicting the gamble students were asked to take in a choice between a chance of infection with COVID-19 and attending class online rather than in-person.

To assess preferences for in-person versus online classes more directly, we asked a simple rating scale question regarding students’ willingness-to-pay for online instruction based upon the average out-of-pocket tuition paid university-wide. (This is about one-third of the tuition charged, and is lower because students tend to receive grants and scholarships.)

“Imagine that you are paying tuition out of pocket, and the annual tuition for online classes is $20,000. All things being equal (you remain in your New York housing and obtain a degree from Columbia University), how much would you be willing to pay if the courses were only available on Zoom as opposed to in-person? Input value between 0 and 20,000.”

Next, we conducted a standard gamble exercise to assess the students’ willingness to expose themselves to infection in a social setting (**Figure 2**).

**Figure 2.**
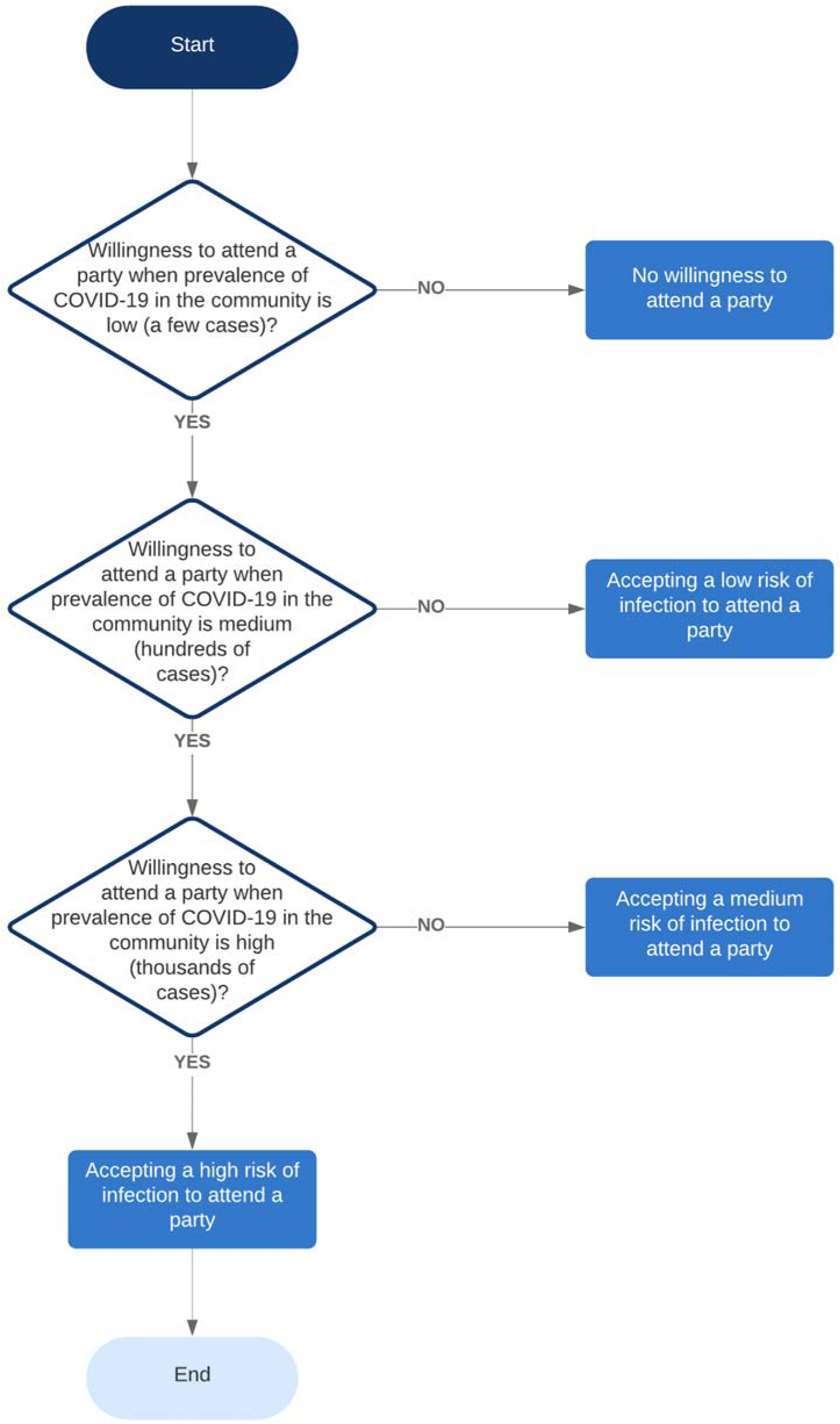
Flow diagram depicting the gamble students were asked to take in choosing to remain at home or to attend a party at different levels of COVID-19 in the community.

“Now imagine that your best friend is planning on having a party in student housing. The circulating rate of COVID-19 in the community is very low at the moment, with just a few people in New York City testing positive. The party will involve drinking and talking with four or five other friends, some of whom you don’t know. Would you be willing to go to the party?”

If the student answered ‘Yes’, then the prevalence of Covid-19 was increased to medium but not negligible (a “few hundred people”) and then to high (a “few thousand people”). At that point, the exercise was stopped.

To assess day-to-day risk tolerance, we asked a simple survey question: “Approximately, how many times a week do you plan to leave the Columbia campus and spend time in the general New York City area, possibly interacting with non-Columbia affiliates (riding the subway, eating at restaurants, etc.)?”

Results are reported as means and standard errors (SE).

## Results

### Sample validation

The mean risk tolerance values and willingness-to-pay values were similar for the initial 30 responses and the final 16 responses (e.g., the two-tailed p-values for risk of infection in classroom, number of times leaving campus per week, and mean willingness-to-pay for tuition were 0.72, 0.55, and 0.70 respectively).

### Desire for in-person instruction

On average, students were willing to accept a 23% (SE = 4%) risk of infection on campus over the semester in exchange for the opportunity to attend class in-person. Of the 46 students, 37 (80%) were willing to accept a >1% chance of infection and 3 (7%) were willing to accept a 100% chance of infection. One student was not willing to attend classes in-person unless the risk was 0%, and 9 (20%) were willing to attend in-person classes if the risk was less than 1%.

With respect to costs, students were willing-to-pay an average of only 48% (SE: 3%) of their tuition if courses were held exclusively online. No student was willing to pay full price for exclusively on-line instruction, and the maximum reported willingness-to-pay for online-only courses was 85% of standard tuition.

### Tolerance for risk of infection in the community

Of the 46 students, 27 (59%) indicated no willingness to attend a party over the entire semester. Of the 19 students who were willing to attend a party with some incidence of COVID-19 in the community, 6(13% of the total sample) indicated that they would do so only if the community prevalence of COVID-19 was low; another 6 (13% of the total sample) were willing to attend in the moderate prevalence scenario, and 7 (15% of the total sample) were willing to attend even if the prevalence of Covid-19 in the community were high (presence of thousands of infectious cases in the community).

Students planned to dine out an average of 3.6 times per week (SE: 0.54). Of the 46 students, 4 (~9%) indicated that they would dine out 10 times or more per week.

## Conclusions

Many universities across the nation opened in the Fall of 2020, while others vacillated between opening, partially opening, and closing.^7^ The Spring of 2021 will require clearer decision making. On average, our sample of students who are knowledgeable about COVID-19-associated risks would be willing to risk a 23% chance of infection in exchange for in-person classes and would be willing to pay only 48% of the cost of in-person classes if were classes held exclusively online. None of the 46 students surveyed were willing to pay full price for online-only courses even assuming they received full academic credit.

However, university and local elected officials alike should be cognizant that this risk tolerance, even when measured among public health students, extends to with more attending large gatherings and to travelling off campus and. About 40% of students of students were willing to attend a party, including over 25% who would attend even if the community prevalence of COVID-19 were moderate or high, and students planned to dine out off-campus an average of 3.6 times per week.

Our study was limited in that it was a sample of 46 recent public health graduates at one school. Moreover, some students reached out to other students to ask them to take the survey. Since no identifiers were collected, it is possible that these additional students had similar risk preferences as the primary respondent. However, the means for the final 16 responses were similar to the initial 30 responses, suggesting that these additional 16 responses produced little influence on the results.

Although our respondents have the requisite knowledge of Covid-19 and had first-hand experience with both online and in-person instruction, our risk assessment should be interpreted with caution. Students who are younger and less knowledgeable may be even more inclined to attend large gatherings.

Finally, we based willingness-to-pay on a round valuation number ($20,000) that was less than the tuition charged by the university ($60,000). We instead used the lower number because it reflects what the average student pays after scholarships and grants.

It is also unclear how students’ risk of infection might differ were classes offered exclusively online and were students to remain off-campus. When classes are held on-campus, students may live in an environment with significantly more infection controls in place than they might have were they to remain in their hometown and take classes online. For example, one work-around for students’ willingness to take on risk is to place students in “cohorts.” In this model, they have opportunities to socialize, but those opportunities are limited to smaller numbers of students who live together and take the same classes together.^8^

University life likely offers more opportunities for socializing than, for example, living with one’s parents. Therefore, it may be that holding classes online-only comes at a lower risk of infection with COVID-19 than holding classes in person.

However, it may be that the “worst-case” scenario for many universities is a situation in which students are invited back to campus only then to switch to online-only instruction. In such cases, students are removed from their hometown, placed in proximity to other students near campus, and then removed from the protective environment of on-campus instruction. In such situations, both student dissatisfaction and the risk of infection may be maximized. For this reason alone, universities with large numbers of students off-campus may wish to: 1) re-consider re-opening or 2) commit to remaining open in the face of surging COVID-19 cases in the Spring of 2021; should they have to close campus, then the “worst-case” situation is likely to be realized.

It is unclear whether the guidelines from the Centers for Disease Control and Prevention (CDC) for re-opening universities will be sufficient to reduce risk on campus.^9^ Our findings suggest that, irrespective of whether they do reduce infection on campus, students appear willing to accept a about a 25% risk of infection in exchange for the opportunity to attend in-person classes, socialize, and dine out. Furthermore, none of the 46 students in our sample was willing to pay full price for online-only instruction, and on average they were willing to pay less than 50% of standard tuition.

This combination of students’ risk tolerance proclivities and their unwillingness to pay tuition places universities in a true conundrum. Some have been willing to assume, the infectious risks, only to shut down because of infectious spread. Others may decide to remain open despite the infectious hazards, perhaps by attempting a variety of “cohort” approaches to reduce risks. Still others are attempting to enrich online-only instruction and to collect most or all of their tuition revenue. Unfortunately, no single instructional model can conclusively balance the desires of students, the usual priorities of universities, and the public’s health.

## Data Availability

The complete dataset is available as a supplementary appendix.

https://www.publichealth.columbia.edu/academics/departments/health-policy-and-management/openup-model

